# How Efficient Can Non-Professional Masks Suppress COVID-19 Pandemic?

**DOI:** 10.1101/2020.05.31.20117986

**Authors:** Yejian Chen, Meng Dong

## Abstract

The coronavirus disease 2019 (COVID-19) pandemic is caused by the severe acute respiratory syndrome coronavirus 2 (SARS-CoV-2), which can be transmitted via respiratory secretions. Since there are currently no specific therapeutics or vaccines available against the SARS-CoV-2, the commen non-pharmaceutical interventions (NPIs) are still the main measures to curb the COVID-19 epidemic. Face mask wearing is one important measure to suppress the pandemic. In order to know how efficient is face mask wearing in reducing the pandemic even with low efficiency non-professional face masks, we exploit physical abstraction to model the non-professional face masks made from cotton woven fabrics and characterize them by a parameter virus penetration rate (VPR) *γ*. Monte Carlo simulations exhibit that the effective reproduction number *R* of COVID-19 or similar pandemics can be approximately reduced by factor *γ*^4^ with respect to the basic reproduction number *R*_0_, if the face masks with 70% *< γ* < 90% are universally applied for the entire network. Furthermore, thought experiments and practical exploitation examples in country-level and city-level are enumerated and discussed to support our discovery in this study and indicate that the outbreak of a COVID-19 like pandemic can be even suppressed by the low efficiency non-professional face masks.

## I. Introduction

The global spread of coronavirus disease 2019 (COVID-19) has already affected over 200 countries and regions within only a few months, and led to more than 300,000 deaths until the middle of May 2020 [1], [2]. The COVID-19 is a pandemic caused by severe acute respiratory syndrome coronavirus 2 (SARS-CoV-2), which can lead to acute respiratory distress syndrome. One main reason for the rapid expansion of this outbreak is the efficient human-to-human transmission [3]. SARS-CoV-2 can be detected in nasal and throat swabs samples obtained from patients indicating high viral loads in upper respiratory tract samples [4]. Therefore, it appears to be likely that virus can be transmitted via respiratory secretions in the form of droplets (> 5*µ*m) or aerosols (< 5*µ*m). It has been reported that the virus can remain active in aerosols for multiple hours [5]. Since there are currently no specific therapeutics or vaccines available against the SARS-CoV-2, the classical public health measures are needed to curb the COVID-19 epidemic. The primary goal of all the measures is to interrupt person-to-person transmission [6]. To accomplish this goal, many common non-pharmaceutical interventions (NPIs) such as isolation and quarantine, social distancing, and community containment have been applied. Except these measures, face mask wearing is emerging as one of the important NPIs for suppressing the pandemic, especially when considering that the pre-symptomatic or asymptomatic cases may also play a critical role in the transmission process [5], [7]. Compared with the societal lockdown, universal masking is far more sustainable than the other measures from economic, social, and mental health standpoints [8]. There are two main types of face masks: the professional mask such as N95 masks and medical masks, which have high efficiency, and the non-professional face masks such as homemade face masks with low efficiency. In our study, we mainly focus on the efficiency of different types of non-professional face masks, since medical masks are in short supply during the COVID-19 pandemic and preferentially used in hospitals not for the public social network. We introduce certain types of cotton face masks which are characterized by their different dimensions of pore diameters, and exploit physical abstraction to model the capability of these face masks to block aerosols. Based on the investigated physical abstraction and parameters, Monte Carlo simulations are carried out to demonstrate the outbreak of COVID-19 pandemic with or without face masks in a social network to check if the low efficiency non-professional face masks manage to slow down the outbreak and spread of COVID-19 or similar pandemics.

## II. Materials and Methods

In this study, abstracting physical and statistical models are our major methodologies for simulating a social network, in which the COVID-19 pandemic starts to be suppressed with the usage of different non-professional face masks. The face mask is modeled as shown in Fig. 1(a). Four different face masks can be characterized by their dimensions of pore diameters. For a given surface area on the face masks, the density of the pores equivalently represents the capability of face masks to block the particles. According to the investigation in [9] for 10 cotton woven fabrics, the pore size varies from 19*µ*m to 115*µ*m. Hence, the face masks, used in this study, are made from cotton woven fabrics, and the corresponding pore size is selected from this range. It is well known that respiratory droplets and aerosols are the major virus carriers. In this study, as illustrated in Fig. 1(b), we focus on the spherical aerosol, which is with even smaller dimension, and can thus be critical during the outbreak of the pandemic. It is reported that the diameter of SARS-COV-2 virus is around 0.08*µ*m to 0.12*µ*m. The median diameter of aerosols is 3.9*µ*m, and 65% of the aerosols have the diameter less than 5.0*µ*m [10]. Thus, we model the diameter *d* of aerosols exactly based on the observations in [10], by means of continuous Poisson distribution [11] with distribution function as

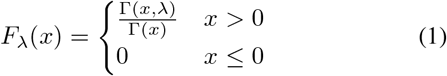

where Γ(*x*, *λ*) denotes the incomplete Euler Γ-function 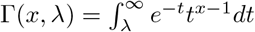 with Γ(*x*) = Γ(*x*, 0) for *x* > 0, λ ≥ 0 and *λ* stands for the mean value of diameter *d*. Furthermore, for a given surface on a face mask, e.g. Fig. 1(a), we assume that the diffusion of SARS-COV-2 aerosols obeys uniform distribution, and the same amount of aerosols approaches the face masks. As illustrated in Fig. 1(b), we assume that aerosols can penetrate the pore of a face mask, if the geometric center of the spherical aerosol locates in the red shadowed region within a pore. We use Monte Carlo simulations to repeat the same scenario for different non-professional face masks and count the number of aerosols and their diameters, which penetrate the pores of the face masks individually. For instance, for face mask *k*, we can compute the sum area of the surfaces of the total penetrated aerosols as

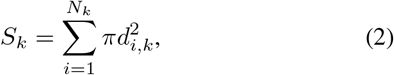

**Figure 1:**
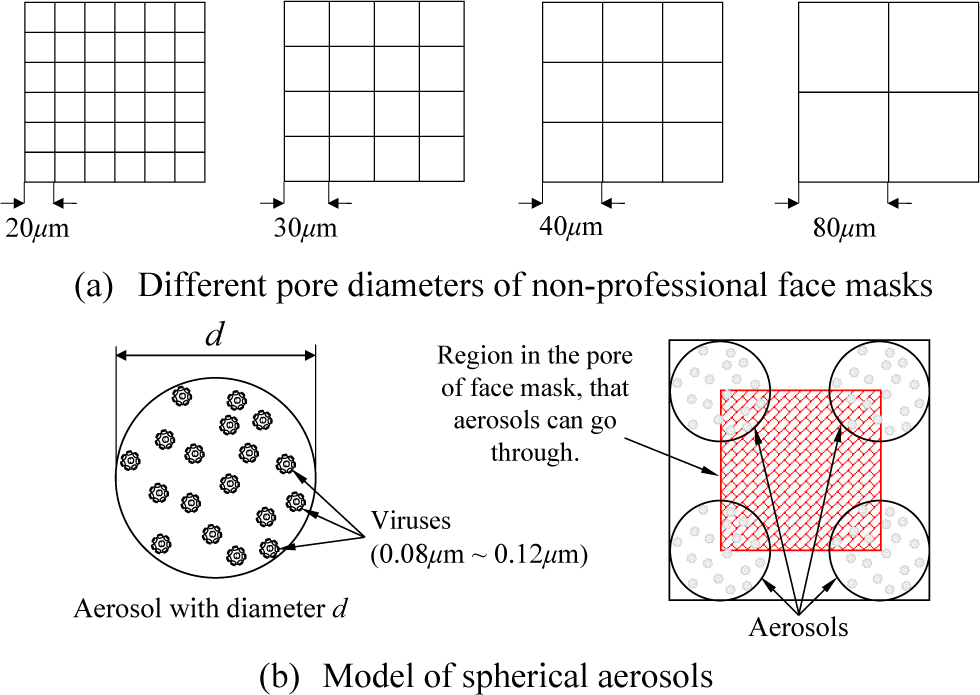
Model of non-professional face masks and aerosols

where *N_k_* stands for the number of penetrate aerosols out of all *N* attacking aerosols, and *d_i,k_* denotes the diameters of these penetrate aerosols. Notice that the sum area *S_k_* represents the amount of the penetrated viruses by deploying face mask *k*, if the density of viruses per unit area on the aerosols is assumed to be constant. Hence, we introduce the virus penetration rate (VPR) *γ_k_* as a ratio between the sum area *S_k_* of penetrated aerosols and the sum area of total approaching aerosols. It holds

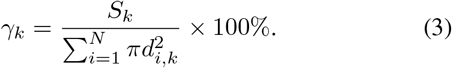

Furthermore, we can similarly define the successful aerosol block rate (ABR) *α_k_* by exploiting face mask *k* as

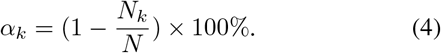

In [12], it is shown that the basic reproduction number of a pandemic *R*_0_ can be formulated as a function

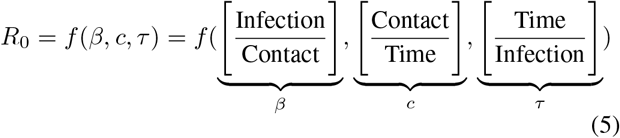

where transmission rate *β* provides the rate of infection of a given contact between a susceptible and infected individual, social contact rate *c* determines the average number of contacts between susceptible and infected individuals and duration of infectiousness is denoted by *τ*. Obviously, face masks can play a role for reducing transmission rate *β* with respect to the VPR *γ_k_*. With face mask *k*, the effective transmission rate *β_k_* can be formulated as

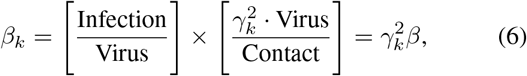

where the amount of viruses per contact is reduced by factor 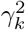, if both susceptible and infected individuals use face masks. Thus, this reduces the rate of infection. Throughout the Monte Carlo simulation in the following section, we assume that there are several options of non-professional face masks within the social network. They are characterized by their VPR *γ_k_* and ABR *α_k_* individually. One of the important motivations of this paper is to demonstrate that even these non-professional face masks can play a significant role to control the pandemic. We will also illustrate how the VPR *γ_k_* of face mask *k* numerically impacts the effective reproduction number *R_k_*.

## III. Results

### A. Characteristics of Face Masks

As shown in Fig. 2, we have a complete overview of VPR *γ_k_* and ABR *α_k_* for the face masks with different pore diameters, which are modeled by equations (1) to (4) through the Monte Carlo simulations. The face masks with pore diameters 20*µ*m ≤ *d_k_* ≤ 120*µ*m are non-professional face masks, consisting of single layer cotton woven fabrics. The corresponding VPR and ABR satisfy 50.71% ≤ *γ_k_* ≤ 90.33% and 6.15% ≤ *α_k_* ≤ 32.92%, respectively. In details, the face mask with pore diameter *d_k_* = 120*µ*m can block 6.15% aerosols and 9.67% viruses, and the face mask with pore diameter *d_k_* = 20*µ*m can block 32.92% aerosols and 49.29% viruses. We notice that the inequality *α_k_* < 1 − *γ_k_* holds, and introduce the compensation factor *θ_k_*, defined as

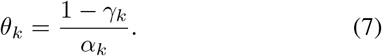

**Figure 2:**
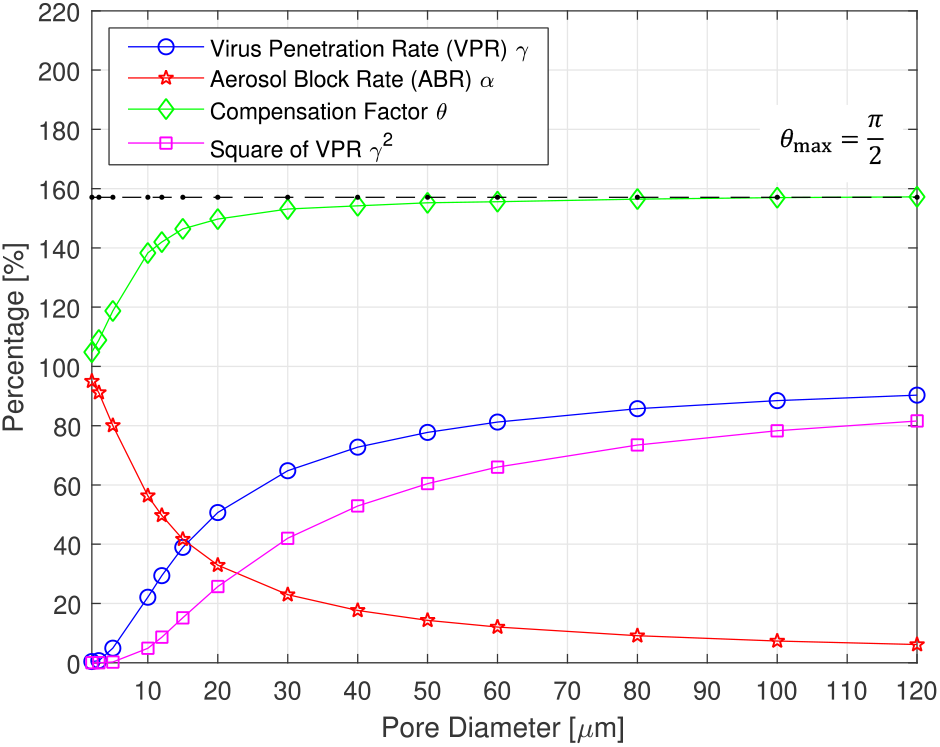
Virus penetration rate (VPR) and aerosol block rate (ABR) of face masks with different pore diameters

The compensation factor *θ_k_* indicates the fact that ABR *α* only counts the number of successfully blocked aerosols. For face masks generally with large pore size, once a successful block happens, the number of the viruses on the aerosol plays a more important weighting in computing *γ_k_* than in *α_k_*. The compensation factor *θ* illustrates that the non-professional face masks with relatively large pore diameters should not be underestimated. Empirically, we observe that the compensation factor *θ* is bounded by 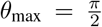, which belongs to a part of future analytical investigation. Finally, we add the effective VPR 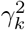 to Fig. 2, if the face mask *k* is exploited by both infectious individuals and susceptible individuals systematically.

### B. Simulate COVID-19 Pandemic in a Social Network

In this section, we study the outbreak of COVID-19 pandemic in a social network by means of Monte Carlo simulation, and reveal different progressing, if the face masks are introduced in the social network. We refer to the COVID-19 pandemic parameters in [13] and [14]. The transmission rate and contact rate are assumed to be *β* = 0.139 and *c* = 6, respectively. The average duration of infectiousness is *τ* = 3 days. In Fig. 3, the outbreak of COVID-19 pandemic in a social network is demonstrated by a one-shot Monte Carlo simulation. With the selected parameters *β, c* and *τ*, the number of daily new infected cases is counted. The infectious transmission, characterized by the generations of the viruses, is also illustrated with different colors. On one side, visualizing the infectious transmission and virus generation development serves as a plausibility check for the simulation. On the other side, Fig. 3 also illustrates that the basic reproduction number can be underestimated at the early stage of the pandemic, as mentioned in [13] and [14] due to lack of data, or as shown in Fig. 3 due to counting only on a one-shot Monte Carlo simulation for the pandemic. Thus, in the following investigation, we repeat such one-shot Monte Carlo simulation as shown in Fig. 3 for 100,000 times and take a final averaging, to get a stationary result for a given pandemic parameter setting. Furthermore, we introduce five types of face masks, which are categorized as 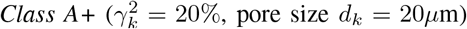, 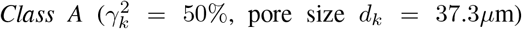, *Class B* 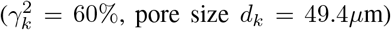, *Class C*, γ^2^_*k*_ = 70% pore size *d_k_* = 70.7*µ*m), and *Class D* 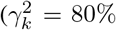, pore size *d_k_* = 110.3*µ*m). The corresponding parameters are obtained from the results in Section III.*A*. In Fig. 4, the accumulated daily number of infections is presented, assuming that one type of face mask out of *Class A+* to *Class D* is systematically exploited in entire social network, without considering the impact from social distancing and hand hygiene [10]. First of all, we focus on the simulation of COVID-19 outbreak in the social network without any measures against the spread of the pandemic. According to the Monte Carlo simulation, the data is fitted in sense of Minimum Mean Square Error (MMSE) criterion between the simulated curve and the target curve. The exponential growth factor is *R*^1*/τ*^_0_ ≈ 1.6957, and thus the basic reproduction number *R*_0_ is approximately 4.8758, which fits to the early analysis in [13] and [14]. The systemic wearing of face masks can effectively slow down spread of the COVID-19 pandemic. By introducing the doubling time *T_d_*, with the assumed average infectious duration *τ*, the corresponding reproduction number of individual face mask can be estimated as

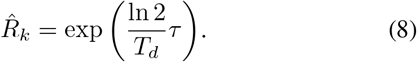

**Figure 3:**
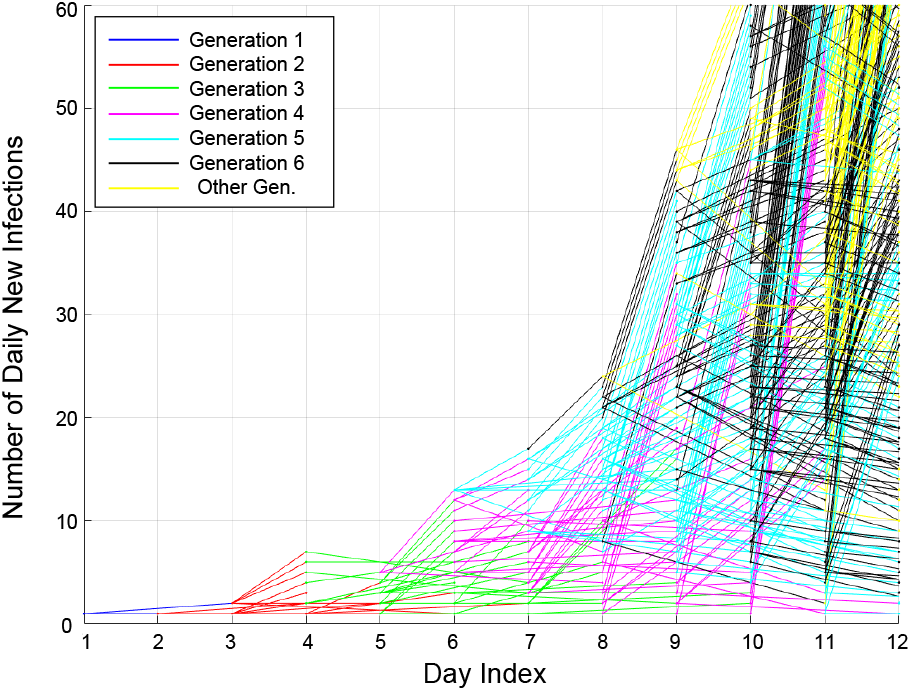
Simulate the outbreak of COVID-19 in a social network: one pandemic realization by one-shot Monte Carlo simulation to visualize the daily new infections and the development of virus generations for given parameters *β, c* and *τ*

In Table I, the reproduction numbers 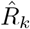 with exploitation of the face masks from *Class A* to *Class D* are estimated based on (8) and summarized. Empirically, we discover the one-to-one correlation of the basic reproduction number *R*_0_ and the reproduction number 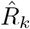 as

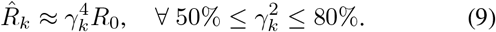

**Table I:**
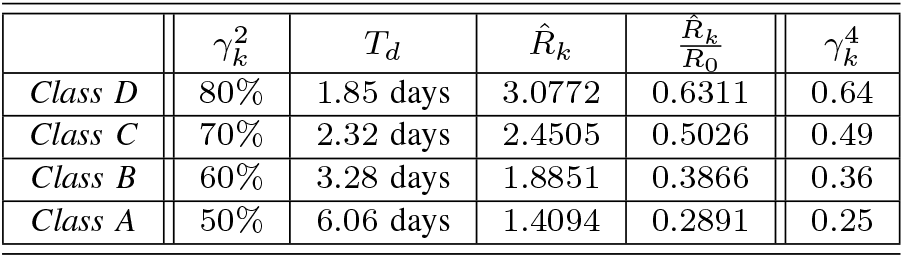
Estimate the Reproduction Number 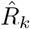 for Face Mask *k*

Equation (9) illustrates the fact that even face masks with large pores can effectively reduce the reproductive index *R*_0_ by a factor of 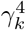. From the Monte Carlo simulation in Fig. 4 with *Class A+* face mask, it is visible that the outbreak curve can be flattened both at the beginning of COVID-19 pandemic or one week after the outbreak, if exploiting the face masks by cotton woven fabric with pore size *d_k_* = 20*µ*m in the entire network. As an extension of Fig. 4, we start the Monte Carlo simulations to study the effect of *Class A+* face masks in the social network to suppress COVID-19 for longer period, when face masks obligation is applied at different stages of the pandemic. Basically, Fig. 5 illustrates the fact that the exploitation of face masks is more effective when exploited in the early stage of pandemic. As shown in Fig. 5, the red curve (*Day 17*) reveals much stronger effect on suppressing the infections than the blue curve (*Day 21*), which exploits the face masks only 4 days later.

**Figure 4:**
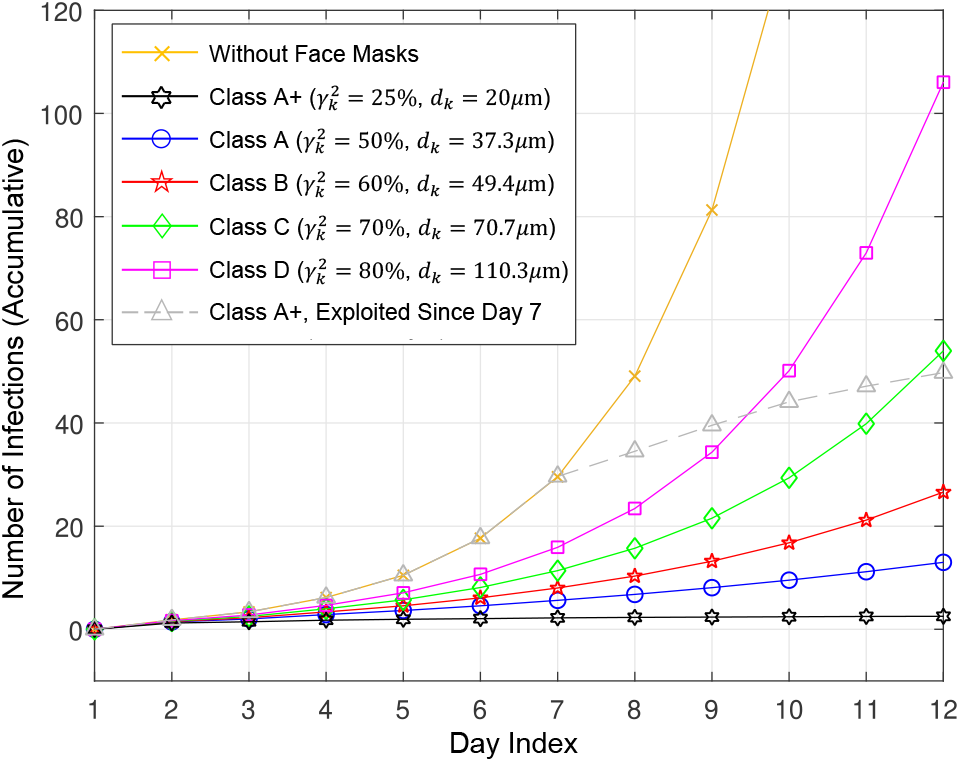
Simulate increasing of daily number of accumulative infections applying diverse face masks: each curve averages the results of 100,000 one-shot Monte Carlo simulations, repeated for given parameters *β, c, τ* and 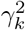

**Figure 5:**
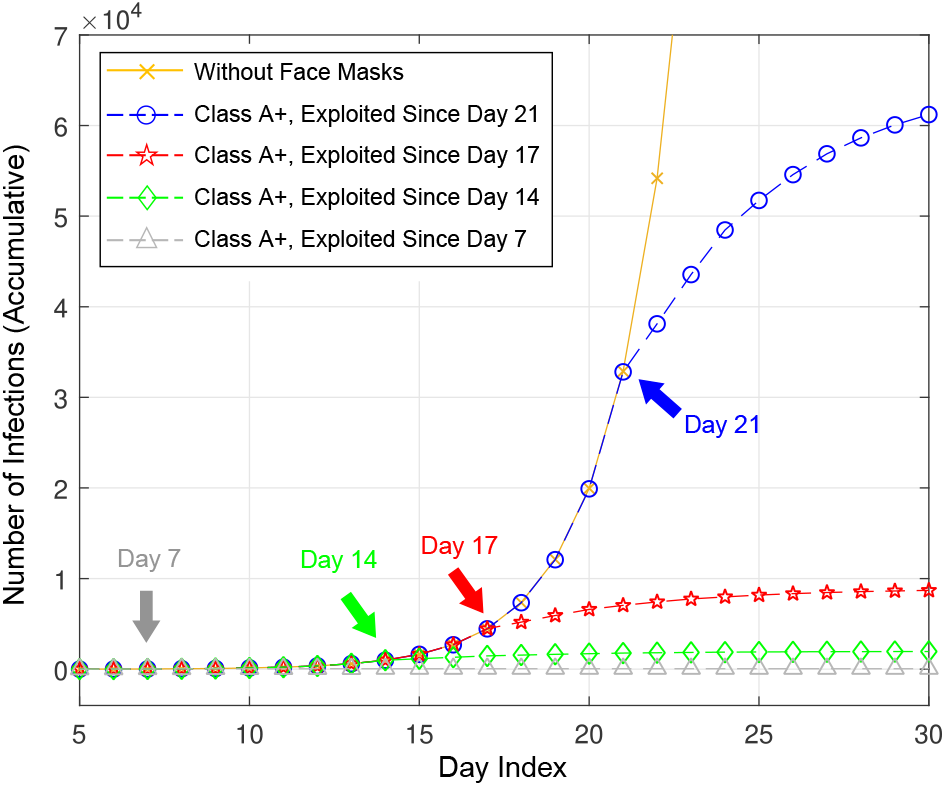
Exploit face masks after the outbreak of COVID-19 pandemic

### C. A Thought Experiment for COVID-19 Pandemic in US

In Fig. 6, the daily increasing of the number of COVID-19 infections in USA is presented. It can be clearly noticed that there are two stages during the early development of the pandemic. In the first stage, the number of infections increases exponentially. This stage is fitted by the red curve considering MMSE criterion. The exponential growth factor can be approximated as 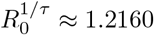. Since the COVID-19 pandemic was relatively underestimated in USA at the early outbreak stage, we select a longer infectiousness duration *τ* = 6 days. Thus, the basic reproduction number in USA is approximately *R*_0_ ≈ 3.233. In the second stage, the development of the pandemic can be linearly fitted. Furthermore, we realize that the green straight line follows the tangent direction at the turning point of the red exponential curve. It is clearly showed that if the period of first stage can hardly be reduced, we should possibly try to flatten the red exponential curve. The usage of face mask can achieve this goal. In Fig. 6, the number of infections will be doubled every *T_d_* = *τ* ln 2*/* ln *R*_0_ ≈ 3.5 days. Let us assume that the social network exploit a face mask with *γ*^2^ = 75%, which is comparable to *Class C* or *Class D* face masks in the Monte Carlo simulations for Fig. 4. The new doubling time of the COVID-19 pandemic in USA is 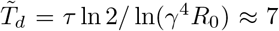 days. Especially, as an important consequence, the slope of the green curve in the second stage will also be reduced, which indicates that the usage of face masks with relatively low efficiency can still be very effective at the early stage of a pandemic.

**Figure 6:**
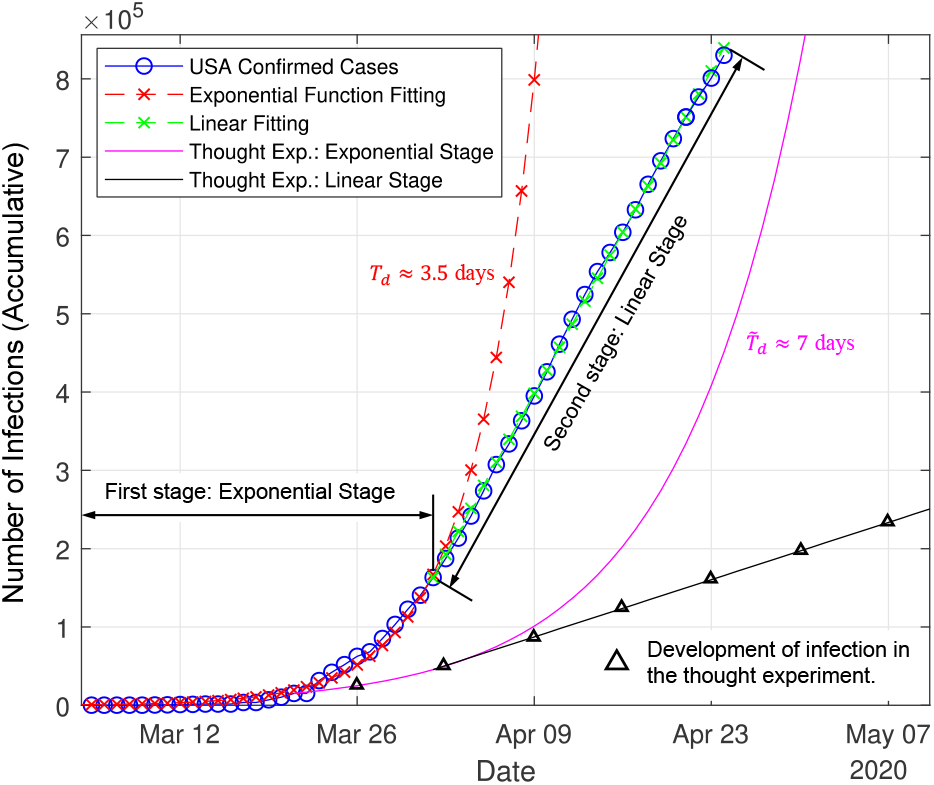
Daily increasing of number of infections in USA [1]

### D. Exploitation of Face Masks in Reality

In previous section, we exploit physical abstractions, mathematical or statistical models, and thought experiments to clarify that even non-professional face masks can help to suppress the propagation of COVID-19 pandemic. In this section, we will verify our models focusing on the development of the pandemic in different countries or cities, which are impacted by exploitation of face masks. In Fig. 7, the development of COVID-19 pandemic in Germany, Austria and Czech are compared to each other. Considering the fairness of the comparison, normalization is introduced, so that the number of infections per 1 Million population is studied for three countries. During the early exponential stage, the reproduction number *R* of Austria was even bigger than that of Germany and Czech. Czech introduced strict mandatory face masks policy on 19 March, which prohibited the movement outside without having mouth and nose covered by a respirator, face mask or similar [15]. The pandemic was effectively controlled. We further compare the development of pandemic in Austria and Germany. After the pandemic stepped into the linear stage, the virus reproduction in Austria was still faster than that of Germany, which coincides with the discussion in the previous section. On 30 March, the Austrian government announced that everyone entering a store had to wear a face mask, effectively since 6 April [16] [17]. And even more strict face masks mandatory policies were introduced in Austria on 14 April. The instantaneous reproduction number of Austria reduced significantly. On 27 April, most German states introduced face masks obligation, which belonged to relatively soft mandatory policies [20]. The daily growth of accumulative infectiousness of Germany behaved similar to Austria before activating strict face masks obligation on 14 April.

**Figure 7:**
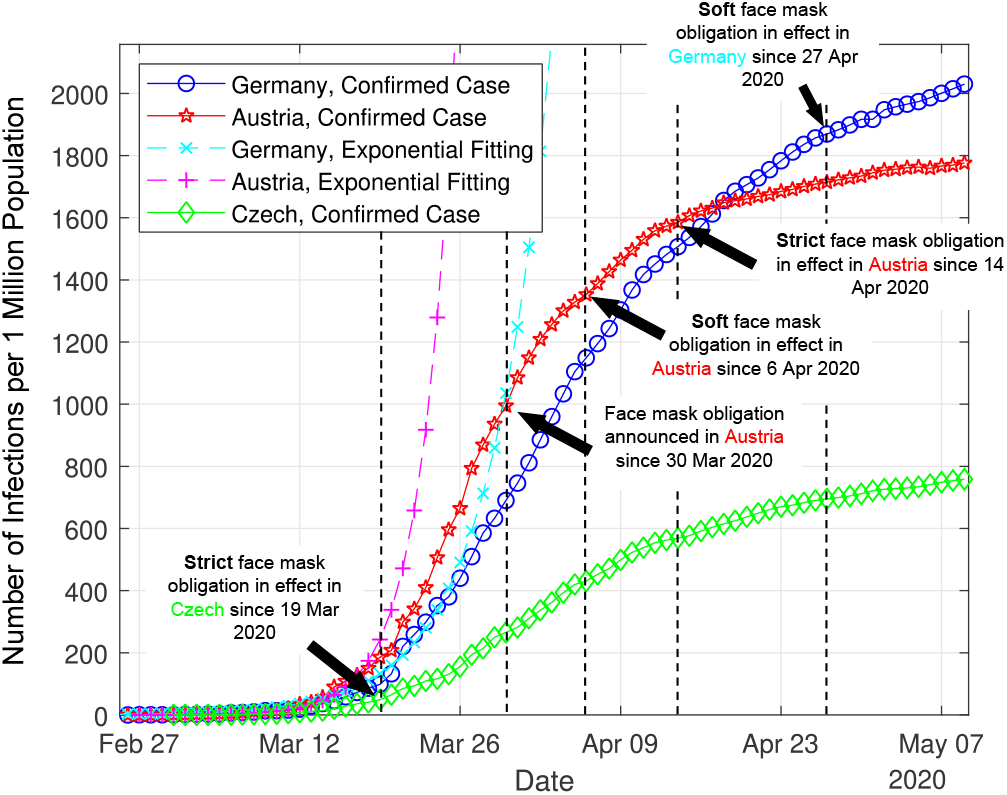
Compare the pandemic in Germany, Austria and Czech [1]

In Fig. 8, the spread of COVID-19 pandemic in different German cities is presented. We select three cities, namely Stuttgart, Ulm and Jena for comparison. Stuttgart is the center of the famous industry region, accommodating 635 thousand population. Ulm and Jena are typical German cities with median scale, with 126 thousand and 110 thousand population, respectively. For the fairness reason, we investigate the number of COVID-19 infection per 100 thousand population for three cities. It can be also observed that situation in Jena seemed to be the worst among three cities at the early outbreak stage of COVID-19 pandemic. On 31 March, Jena was the first German city to announce an obligation to wear masks, or makeshift masks including scarves, in supermarkets, public transport, and buildings with public traffic [20], [21] and the policy was in effect on 6 Apr. With the mandatory face masks policy, the daily infection number of Jena decreased dramatically and reached to 0 for complete 13 days until 22 April. Although Stuttgart and Ulm adopted similar policies except the obligation for face masks, the number of infections could not be reduced during the same time period from 9 April to 22 April. On 27 April, Stuttgart and Ulm started the mandatory face masks policy. The curve of number of infectiousness started to becoming flattened.

**Figure 8:**
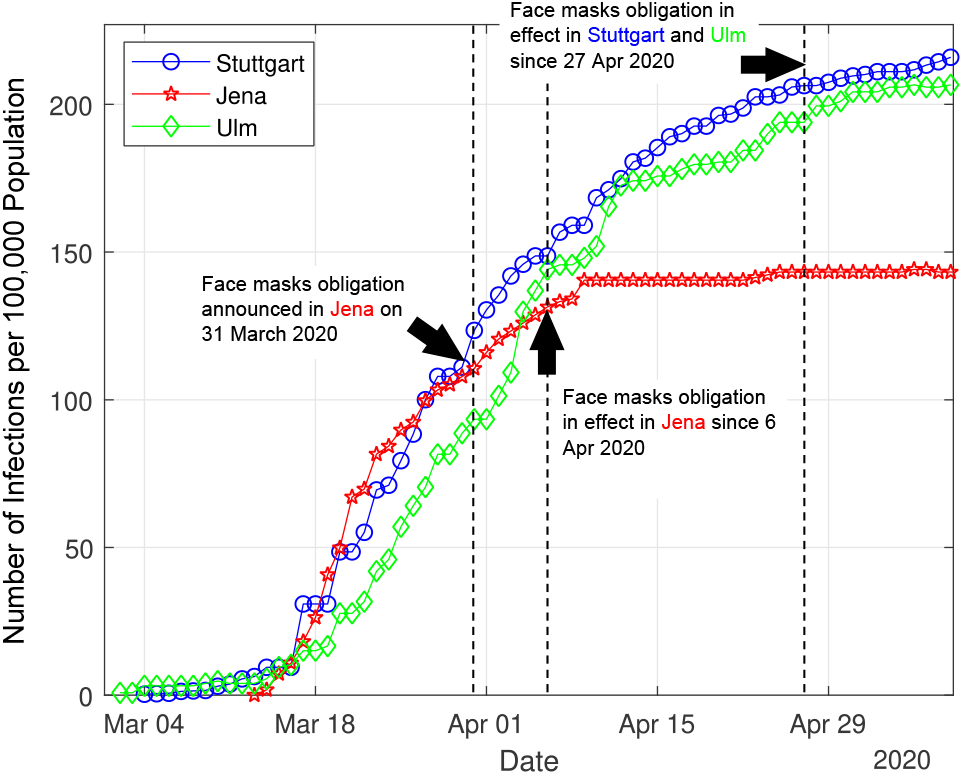
Compare the pandemic of German cities [18] [19]

## IV. DISCUSSION

In this study, we exploit physical abstraction, statistical and numerical methods to illustrate the important features and the corresponding behaviors of face masks to successfully slow down the outbreak of COVID-19 or similar pandemic. With Monte Carlo simulations, it is numerically demonstrated that even the non-professional face masks can significantly impact the pandemic, if they are systematically deployed in the entire social network. With the example of current development of COVID-19 pandemic in USA, we demonstrate that the outbreak of COVID-19, in sense of the increasing of infection numbers in the social network, consists of an early exponential increasing stage and a linear increasing stage afterwards. It is analytically shown that the speed of the reproduction of the infectiousness in the network can be effectively slowed down, if face masks are applied in the exponential stage. Especially, the reduced reproduction in exponential increasing stage can yield consequently a linear increasing stage with reduced infectiousness reproduction, which will be meaningful. Finally, we explore the data from the reality to compare the outbreaks of COVID-19 pandemic in different locations in country-level or city-level. The results clearly prove the finding obtained in our study and lead to the final conclusion: Face mask wearing is one essential measure to suppress the COVID-19 pandemic. Even the low efficiency non-professional face masks can reduce the virus transmission. Since face mask wearing is far more sustainable than the other measures, it should be applied strictly and universally in the social network during the COVID-19 pandemic period.

## Data Availability

All relevant data are available via the link https://pan.baidu.com/share/init?surl=aPAdSQOnyif-T9-cujJxjA

